# Durable reprogramming of neutralising antibody responses following breakthrough Omicron infection

**DOI:** 10.1101/2023.02.19.23286159

**Authors:** Wen Shi Lee, Hyon-Xhi Tan, Arnold Reynaldi, Robyn Esterbauer, Marios Koutsakos, Julie Nguyen, Thakshila Amarasena, Helen E Kent, Anupriya Aggarwal, Stuart G Turville, George Taiaroa, Paul Kinsella, Kwee Chin Liew, Thomas Tran, Deborah A Williamson, Deborah Cromer, Miles P Davenport, Stephen J Kent, Jennifer A Juno, David S Khoury, Adam K Wheatley

## Abstract

SARS-CoV-2 breakthrough infection of vaccinated individuals is increasingly common with the circulation of highly immune evasive and transmissible Omicron variants. Here, we report the dynamics and durability of recalled spike-specific humoral immunity following BA.1 or BA.2 breakthrough infection, with longitudinal sampling up to 8 months post-infection. Both BA.1 and BA.2 infection robustly boosted neutralisation activity against the infecting strain while expanding breadth against other Omicron strains. Cross-reactive memory B cells against both ancestral and Omicron spike were predominantly expanded by infection, with limited recruitment of *de novo* Omicron-specific B cells or antibodies. Modelling of neutralisation titres predicts that protection from symptomatic reinfection against antigenically similar strains will be remarkably durable, but is undermined by novel emerging strains with further neutralisation escape.

**One sentence summary:** Omicron breakthrough infection elicits durable neutralising activity by recalling cross-reactive vaccine-elicited memory B cells.

## Introduction

SARS-CoV-2 continues to cause significant morbidity and mortality. While currently licensed vaccines based on the ancestral strain of SARS-CoV-2 (Wuhan-Hu-1) are effective at preventing severe COVID-19, they have reduced effectiveness at preventing infection with novel variants that escape vaccine-elicited neutralising antibodies. The Omicron variant is highly antigenically distinct and rapidly outcompeted the Delta variant to become the dominant global strain in early 2022. Various sub-lineages of Omicron (BA.1, BA.2, BA.4, BA.5 and others), each with different degrees of antibody evasion, have since emerged in successive waves (*1, 2*). The high transmissibility and immune evasion of Omicron, in tandem with waning of vaccine-elicited immunity, have resulted in increasing frequencies of “breakthrough” infections of vaccinated individuals. At a population level, immunity against SARS-CoV-2 is thus becoming increasingly complex, with variable dosing and types of vaccines, infection with distinct variants, or a combination of both (hybrid immunity).

Recent studies of antibody and memory B cell responses following breakthrough infection with Delta or Omicron BA.1 have established rapid anamnestic recall of spike-specific antibody responses, reactivation of spike-specific memory B cells and differentiation of antibody-secreting cells (*3-5*). Breakthrough Omicron infection in individuals with 2 prior vaccine doses has also been associated with increases in the breadth of serum neutralising antibody activity compared to those receiving a third dose of vaccine (*3, 6*). Increased neutralising breadth could be derived from *de novo* antibody responses against neo-epitopes within Omicron spike, or alternatively the selective re-expansion of cross-reactive memory B cells established during vaccination. Here we intensively examined the early kinetics of recalled immunity following Omicron BA.1 or BA.2 breakthrough infections, as well as profiled the durable changes in serological neutralisation breadth following recovery. We find that breakthrough infections, despite mild disease course, were highly efficient at both recalling spike-specific memory responses established by prior immunisation, as well as generating novel responses to the viral nucleocapsid. While BA.1 or BA.2 neutralising titres were low or undetectable at early timepoints following infection, these responses expanded robustly and demonstrated breadth against the more escaped BA.4 variant. Longitudinal follow up revealed remarkably stable maintenance of Omicron-specific neutralising activity, which we then modelled to estimate the protective window against reinfection with the same or novel variants with further immune escape. Understanding the impact of periodic infection with increasingly distinct SARS-CoV-2 variants upon the durability and breadth of antibody and memory B cell immunity will be critical to informing optimal design and deployment of COVID-19 vaccines to maximise population-level protection against future variants.

## Results

### BA.1 or BA.2 breakthrough infection drives high viral loads and is immunogenic despite mild disease course

Twenty-six vaccinated individuals (3 with two prior vaccines, 21 with three prior vaccines and 2 previously infected with ancestral virus and subsequently vaccinated) were recruited following Omicron breakthrough infection that occurred at a median of 95 days (IQR 78-124) after receiving their last COVID vaccine dose (Table S1). All individuals reported a mild but symptomatic disease course. Omicron BA.1 infection was confirmed using whole genome sequencing for 8 individuals, and BA.2 confirmed for 15 individuals, with the remaining presumptively designated BA.1 (2 individuals) or BA.2 (1 individual) infections based on the prevalent variants circulating at the time. Serial blood samples and nasal swabs were obtained up to 247 days of follow up, with intensive sampling during the acute infection phase (Figure 1A).

**Figure 1.**
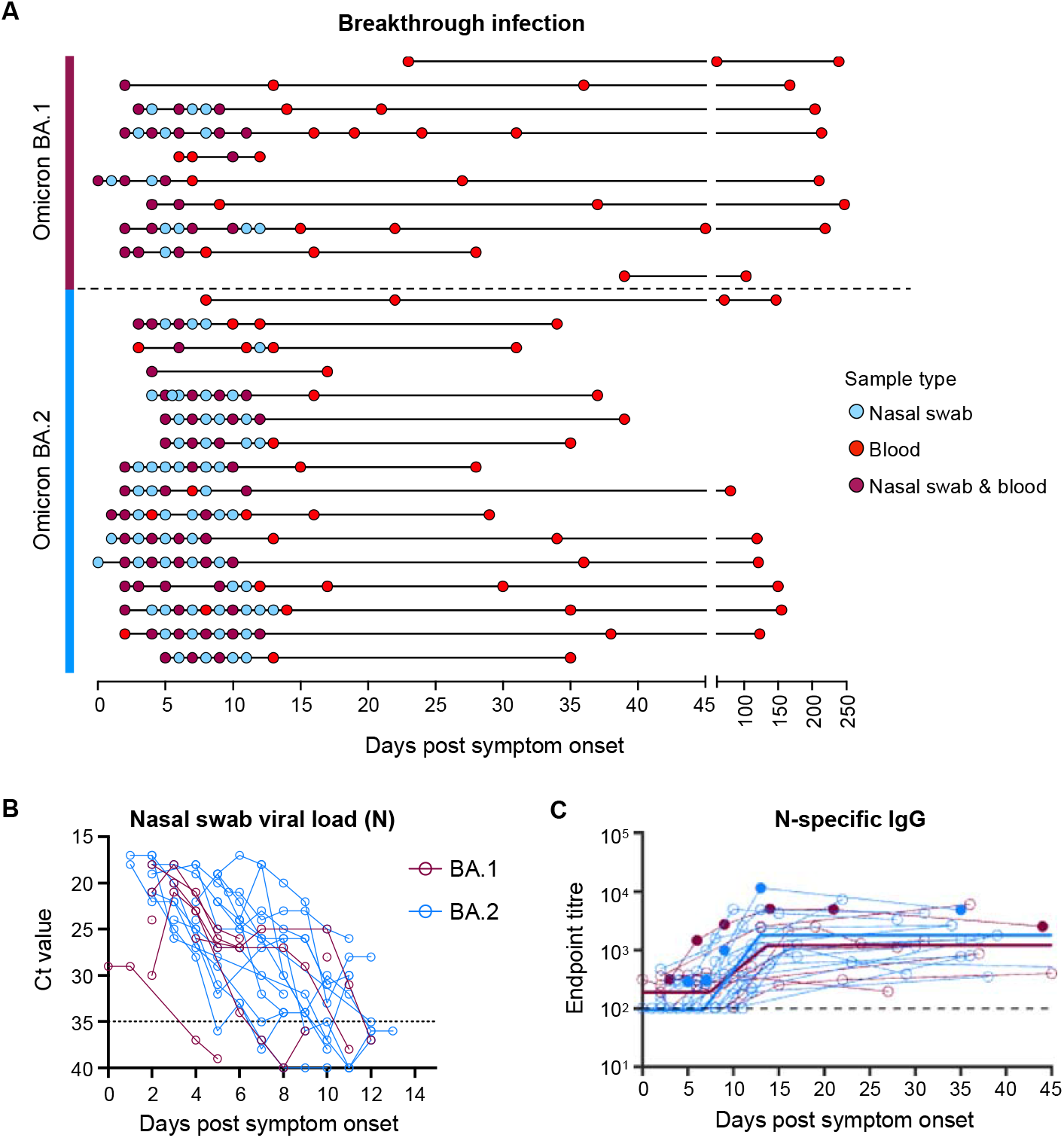
Viral load kinetics and seroconversion to N following Omicron BA.1 and BA.2 breakthrough infection. **(A)** Schematic of longitudinal sample collection following breakthrough infection of vaccinated individuals with Omicron BA.1 (n=10) or BA.2 (n=16). Each line represents a single subject, and each point represents a sample collection (blue, nasal swab; red, blood; purple, both nasal swab and blood). **(B)** Kinetics of SARS-CoV-2 viral load in nasal swabs measured by qPCR of the nucleocapsid (N) gene. **(C)** Kinetics of plasma IgG titres against SARS-CoV-2 nucleocapsid (N) following breakthrough infection with BA.1 (red) or BA.2 (blue). Subjects with previous SARS-CoV-2 infection are depicted in closed circles. The thick lines represent the mean estimate from the piecewise linear regression model using the estimated parameters.

SARS-CoV-2 viral load in nasal swabs was measured via qPCR of the nucleocapsid (N) gene (Figure 1B). Omicron BA.1 and BA.2 breakthrough infection showed similar viral kinetics, with most individuals displaying peak viral load upon recruitment at around 1-3 days post-symptom onset. Median peak Ct values were consistent with previous reports demonstrating robust viral replication during Omicron breakthrough infection despite prior immunisation (*7, 8*). 19 of 21 individuals in our cohort had no prior documented COVID-19 infection and were immunised with COVID-19 vaccines encoding only the spike antigen (BNT162b2, ChAdOx1 nCoV-19, mRNA-1273, NVX-CoV2373) (Table S1). Supporting this, serological responses against SARS-CoV-2 N were at low or undetectable levels at the earliest timepoints sampled (Figure 1C).

However, a clear and consistent expansion of N-specific IgG was observed in plasma samples from ∼7 days post-symptom onset, with similar trajectories for both BA.1 and BA.2 breakthrough infection (Table S2). Overall, individuals with Omicron breakthrough infection in our cohort exhibited marked viral replication in the upper respiratory tract and seroconverted to N.

### Durable boosting of neutralising antibody responses following BA.1 or BA.2 breakthrough infection

At the earliest sampled timepoint following infection (<5 days post-symptom onset), the majority of subjects had robust serological neutralisation activity against ancestral SARS-CoV-2 virus (VIC01), but low or undetectable neutralisation titres against BA.1 or BA.2 (Figure 2A). Neutralising titres against the ancestral strain were rapidly recalled by infection, starting from day 3-4 onwards and concomitant with rises in neutralisation activity against the matched infecting strain. One BA.1-infected individual (CP69) had a transient rise in BA.1 neutralisation activity from days 9-21, before waning to undetectable levels at day 44 post-symptom onset. However, in all other subjects, recovery from infection was associated with robust boosting of BA.1 or BA.2 plasma neutralisation activity.

**Figure 2.**
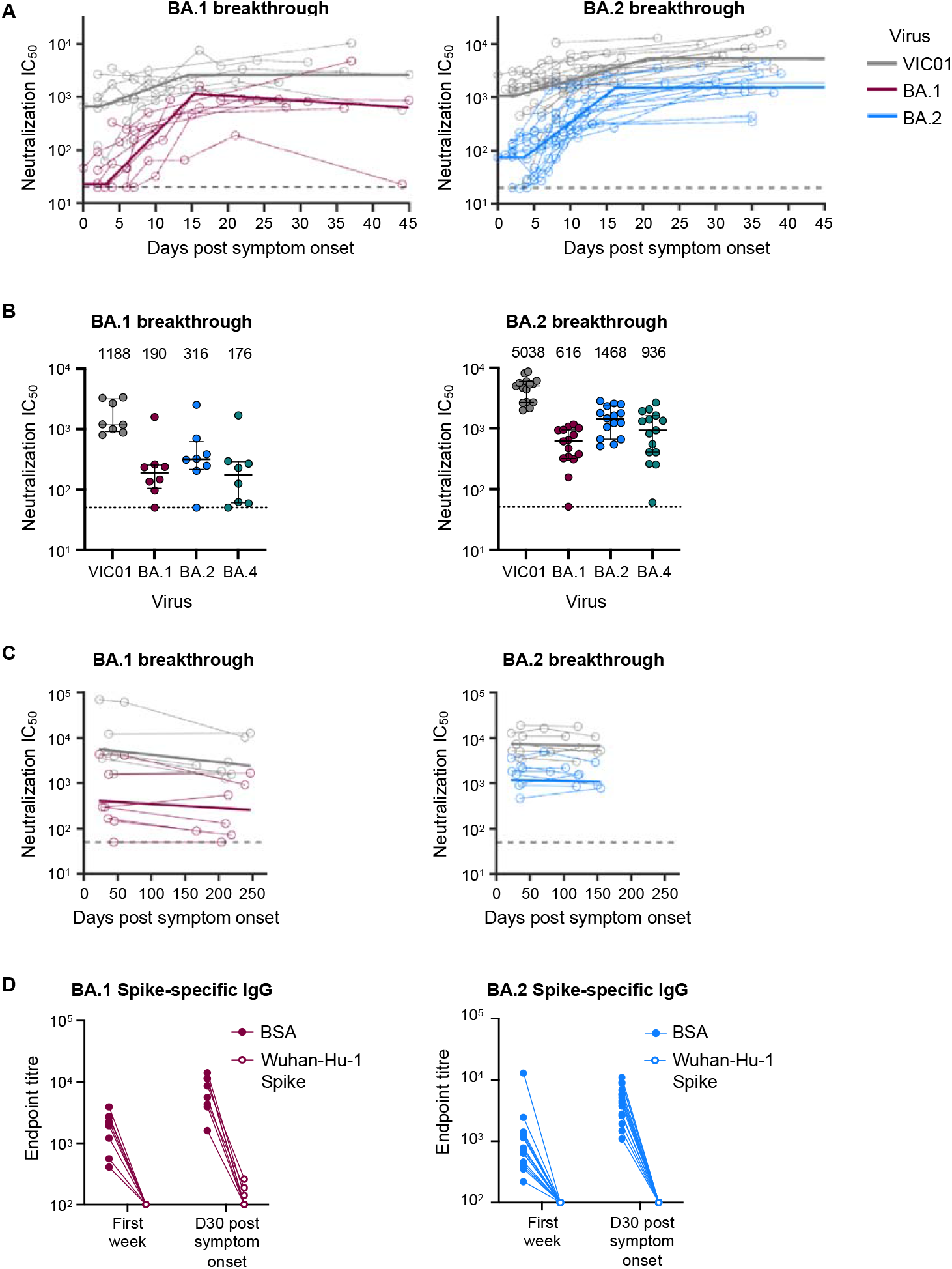
Omicron BA.1 and BA.2 breakthrough infection rapidly recalls neutralising antibodies that are broad and durable. **(A)** Kinetics of plasma neutralisation activity following breakthrough infection against ancestral VIC01 or matched infecting Omicron BA.1 and BA.2 strains. Thick lines represent the mean estimate from the piecewise linear regression model using the estimated parameters. Plasma neutralisation activity was measured using a live virus neutralisation assay against SARS-CoV-2 clinical isolates in HEK293T cells transduced with ACE2 and TMPRSS2. **(B)** Neutralisation mediated by BA.1 and BA.2 breakthrough plasma against ancestral VIC01, Omicron BA.1, BA.2 and BA.4 strains at a median of 34 days post-symptom onset. Data are presented as median ± IQR. **(C)** Longitudinal decay kinetics of plasma neutralisation activity following breakthrough infection against ancestral VIC01 or matched infecting Omicron BA.1 or BA.2 strains up to 4-7 months post-symptom onset. The best fit decay slopes (thick lines) are depicted. **(D)** IgG antibody endpoint titres against BA.1 spike for BA.1 breakthrough subjects (red) and against BA.2 spike for BA.2 breakthrough subjects (blue) following pre-incubation with BSA control (closed circles) or ancestral Wuhan-hu-1 spike (open circles).

We applied a piecewise linear regression model to parameterise the kinetics of antibody recall, including estimates of the initial period of delay, the rate of increase, and time to maximal titres (Table S3). The initial delay phase before detectable increases in neutralising activity against the homologous infecting strain was similar between BA.1 or BA.2 infected subjects, at 3.1 and 3.6 days respectively, with a doubling time of 2.1 and 2.8 days thereafter. Robust expansion of BA.1 and BA.2 neutralising activity against the infecting homologous strains was observed in both BA.1 and BA.2 infected cohorts (31- and 34.7-fold respectively), contrasting with the more modest rise of neutralising activity against VIC01 (5.4- and 15.6-fold respectively).

We next examined the breadth of neutralising antibody responses following breakthrough infection with a panel of Omicron variants including the more immune evasive BA.4 variant. At around 1-month post-symptom onset (median 34 days, IQR 28.5-36.5), individuals with BA.1 breakthrough infection had median neutralisation titres of 1188 against VIC01, 190 against BA.1, 316 against BA.2 and 176 against BA.4 while individuals with BA.2 breakthrough infection had median titres of 5038 against VIC01, 616 against BA.1, 1468 against BA.2 and 936 against BA.4 (Figure 2B). The degree of escape relative to ancestral virus was consistent between the two cohorts, at ∼6-fold for BA.1, ∼4-fold for BA.2 and ∼6-fold for BA.4. Neutralising titres were consistently higher in subjects with BA.2 breakthrough infection, however this is likely reflective of higher baseline titres in these individuals rather than any differential immunogenicity between BA.1 and BA.2.

The longevity of neutralising antibody responses following breakthrough infection was examined in a subset of BA.1-infected (n=7, days 167-247) and BA.2-infected subjects (n=8, days 80-155) (Table S1). After BA.1 infection, BA.1 neutralising titres were stably maintained and readily detectable 5-8 months post-infection, with a half-life of 334 days (Figure 2C, Table S4). An exception was a single subject (CP69) who did not have detectable titres after day 44. Neutralisation against ancestral VIC01 decayed at a half-life of 183 days and remained high (>1:1000) for all individuals at the late timepoint. Durable BA.2-specific neutralising responses were similarly observed in BA.2-infected subjects, with a half-life of 1050 days. Notably, 4 out of 8 BA.2-infected individuals exhibited an increase in neutralising activity between early and late timepoints, indicating that peak titres may have occurred more than 1-month post-symptom onset (Figure 2C, Table S4). Neutralisation against ancestral VIC01 virus was comparably durable and maintained over the 3–4-month period, with a half-life of 1216 days.

### Omicron BA.1 and BA.2 breakthrough infection recalls antibodies that are predominantly cross-reactive against ancestral SARS-CoV-2 spike

Robust and broad neutralising responses against Omicron strains following breakthrough infection could derive from recalled immunological memory or comprise *de novo* BA.1 or BA.2-specific antibodies. We pre-incubated plasma samples with Wuhan-Hu-1 spike or a BSA control to sequester antibodies that bind to ancestral spike, and probed for residual binding activity against BA.1 or BA.2 spike by IgG ELISA. In plasma samples taken during the first week post-symptom onset, pre-incubation with ancestral spike eliminated any residual biding to the BA.1 and BA.2 spike, with no evidence for omicron-specific reactivity (Figure 2D). However, after recovery (∼1-month post-symptom onset), low titres of antibodies specific for BA.1 alone were detected in 3 of 8 individuals. In contrast, no BA.2-infected subjects had antibodies specific for BA.2 alone. This demonstrates that antibody responses to BA.1 and BA.2 breakthrough infection is dominated by cross-reactive specificities that also bind to ancestral spike.

### Omicron BA.1 and BA.2 breakthrough infection primarily recalls memory B cells cross-reactive with ancestral SARS-CoV-2 spike

In individuals with prior SARS-CoV-2 immunity established by ancestral spike vaccines, Omicron breakthrough infection could potentially broaden the memory B cell repertoire by eliciting B cells against neo-epitopes within Omicron spike. We therefore examined the recall of cross-reactive spike-specific memory B cells established primarily through prior vaccination or the elicitation of *de novo* Omicron-specific memory B cells. Spike-specific memory B cells within PBMC samples were stained using a combination of fluorescent spike probes (ancestral Wuhan Hu-1 spike in combination with either BA.1 or BA.2 spike; gating in Figure S1). We observed substantial expansion of class-switched memory B cells (CD19+ IgD-) that were cross-reactive to SARS-CoV-2 ancestral spike and BA.1/BA.2 spike (Figure 3A). Frequencies of cross-reactive memory B cells peaked around 1-month post-symptom onset, before subsequently waning over 100 days of follow up (Figure 3B). We did not observe notable expansion of monospecific memory B cells against either BA.1 or BA.2 Spike (Figure 3A). BA.1 and BA.2 monospecific memory B cells were on average 7 and 16-fold lower than their cross-reactive counterparts around 1-month post-symptom onset, with frequencies remaining unchanged throughout the longitudinal sampling period (Figure 3B).

**Figure 3.**
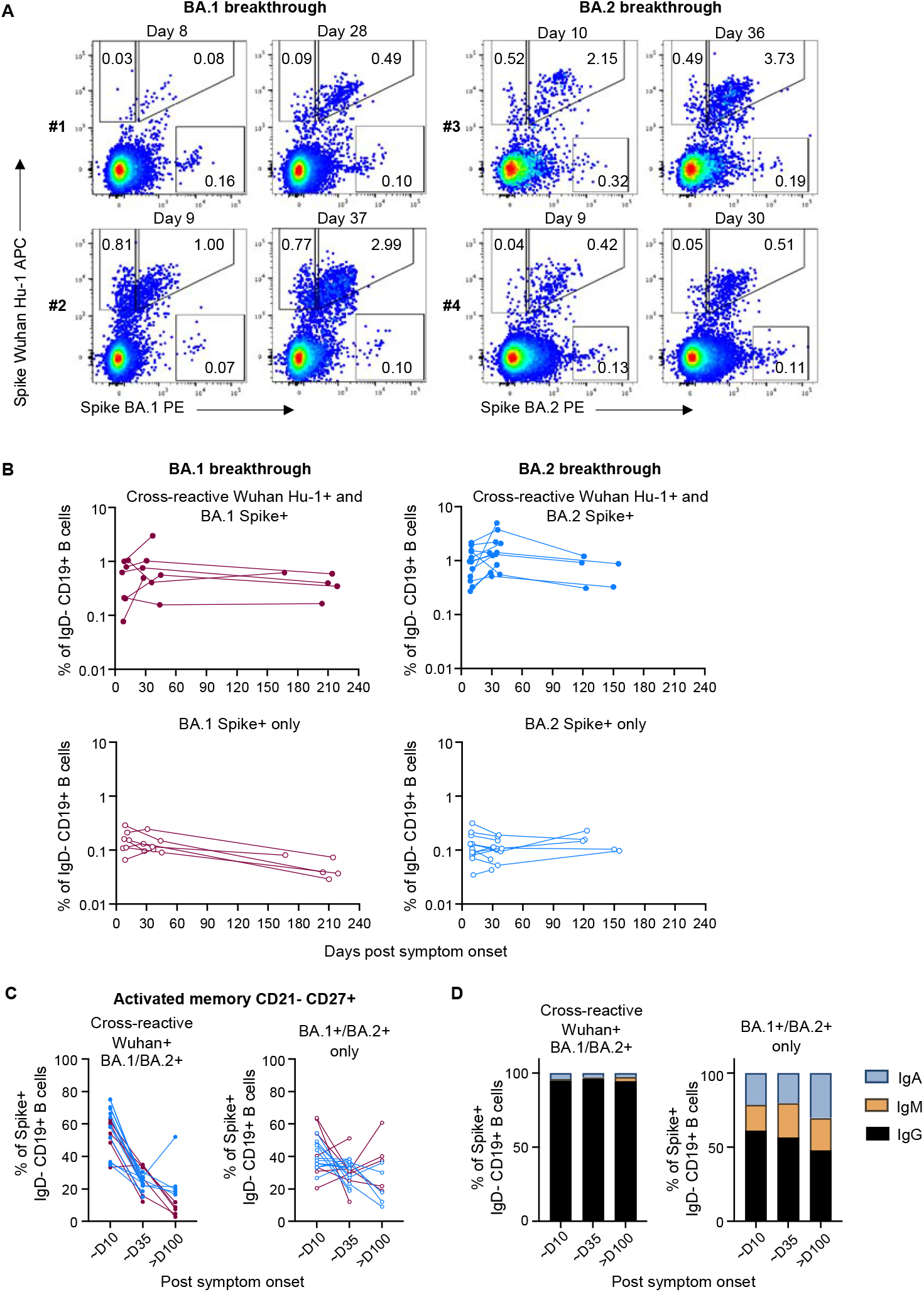
Omicron BA.1 and BA.2 breakthrough infection primarily recalls memory B cells that are cross-reactive against ancestral spike. **(A)** Representative flow cytometry plots of memory B cells (CD19+ IgD-) stained with ancestral (Wuhan Hu-1) and BA.1 or BA.2 fluorescent spike probes, from two individuals with BA.1 breakthrough infection and two with BA.2 breakthrough infection at around day 10 and 30 post-symptom onset. **(B)** Frequencies of memory B cells (CD19+ IgD-) from BA.1 breakthrough subjects (red) and BA.2 breakthrough subjects (blue) that are cross-reactive with Wuhan Hu-1 and BA.1/BA.2 spike (top) or memory B cells that only recognise BA.1/BA.2 spike (bottom) over time. **(C)** Frequencies of activated memory B cells (CD21-CD27+) that are cross-reactive or specific for BA.1/BA.2 spike from BA.1 breakthrough subjects (red) and BA.2 breakthrough subjects (blue). **(D)** Antibody isotype distribution of cross-reactive or BA.1/BA.2 spike-monospecific memory B cells.

A large proportion of cross-reactive memory B cells exhibited an activated phenotype (CD21-CD27+) in PBMC samples taken ∼day 10 post-symptom onset (median 58%), indicating efficient antigen recall during Omicron breakthrough infection, which later waned over the course of recovery (median 14% at ∼day 160) (Figure 3C). We observed a distinct lack of activation of BA.1 or BA.2 monospecific memory B cells, suggesting these cells likely represent background binding or B cells with low affinity for spike (Figure 3C). Cross-reactive memory B cells were predominantly IgG+ at all timepoints, with minor subpopulations of IgA+ or IgM+ isotypes (Figure 3D). In contrast, 45-60% of BA.1/BA.2 monospecific memory B cells were IgG+, which remained relatively unchanged after breakthrough infection, suggesting limited exposure to antigen in vivo. Overall, our data suggests that the cross-reactive memory B cells predominates the recall response, with Omicron-specific populations remaining unexpanded, following Omicron breakthrough infection.

### Estimated protection against reinfection following breakthrough infection is robust but moderated by immune escape

The immunity gained at a population level via breakthrough infection remains unclear. Using the previously developed model for predicting protection from symptomatic infection based on neutralisation titres (*9*), we estimated the durability of protection following Omicron breakthrough infection (Figure 4A, 4B). Since decay rates of neutralising antibodies against the infecting strains were not significantly different across BA.1 or BA.2 breakthrough infection, nor across different variants (ancestral vs Omicron) (Table S4), we pooled neutralisation data for all subjects across all variants and fitted a linear mixed effects model to estimate an average decay rate of 880 days. Using the geometric mean peak neutralisation titres from BA.1 and BA.2 breakthrough infection and the global decay rate, we predict that 70% protective efficacy against the homologous infecting strain would last approximately 4.5 years for BA.1 breakthrough and 7 years for BA.2 breakthrough. Similarly durable protective efficacy was estimated for the ancestral variant, lasting more than 8.5 and 10 years above 70% for BA.1 and BA.2 breakthrough respectively. Forecasting protection to a variant to which participants had not been exposed (e.g., BA.4), protective efficacy was predicted to be maintained above 70% for 705 and 1607 days for BA.1 and BA.2 breakthrough respectively.

**Figure 4.**
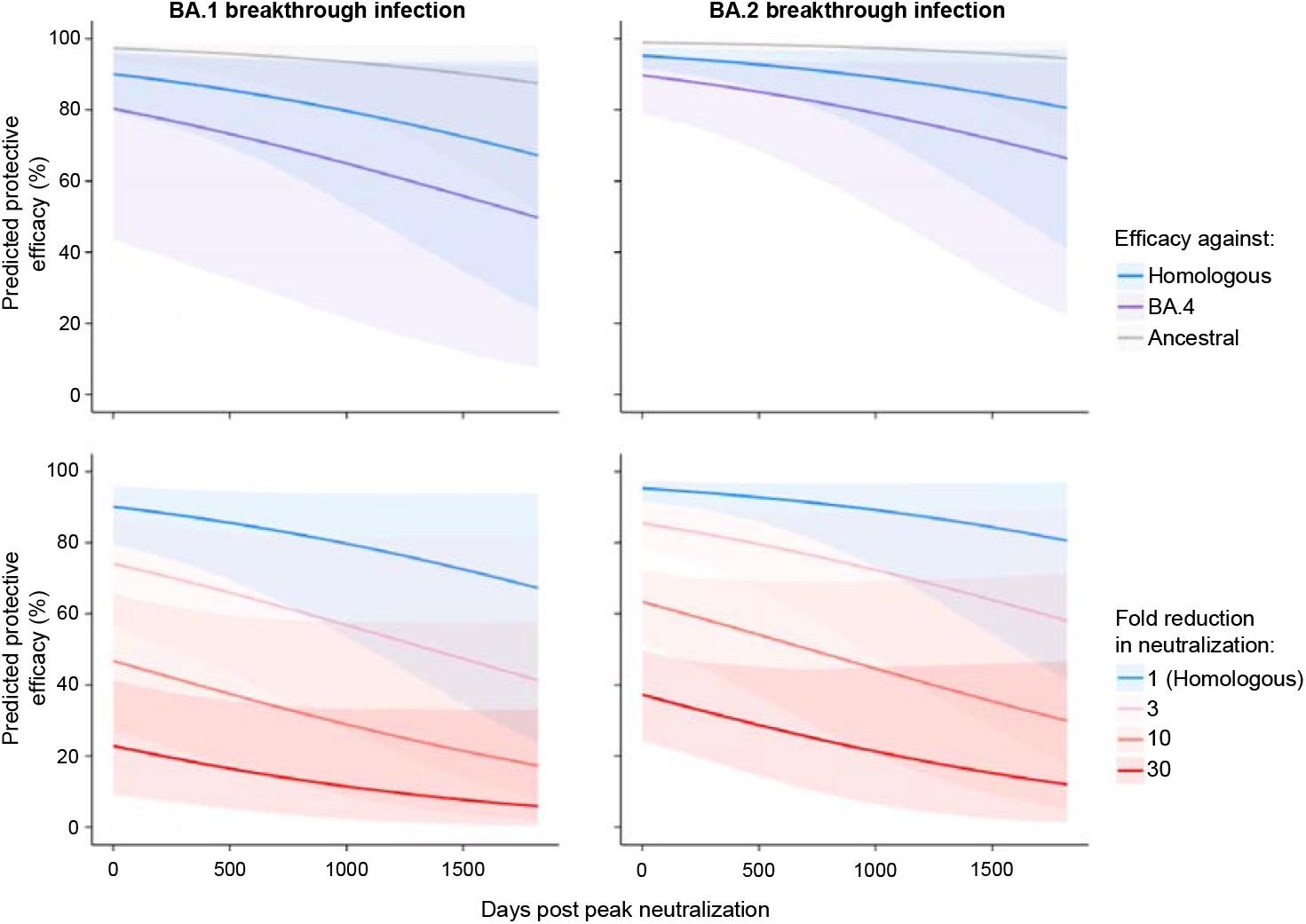
Modelling the protective efficacy against symptomatic SARS-CoV-2 re-infection following BA.1 and BA.2 breakthrough infection. **(A, B)** The predicted decay in efficacy against symptomatic re-infection after peak neutralisation (about day 30 post-symptom onset) based on the peak neutralisation titres observed against ancestral virus (grey), the homologous infecting Omicron variants (blue) and Omicron BA.4 (purple; which individuals had not yet encountered) using the aggregate decay rate estimated. **(C, D)** The predicted decay in efficacy after peak neutralisation titres given a theoretical loss of neutralisation (i.e. 1, 3, 10, or 30-fold loss of neutralisation) to a new variant compared to the homologous neutralisation titres after breakthrough infection (i.e. blue lines in A, B). Lines are predicted efficacies and shaded regions indicate 95% confidence intervals of predicted efficacies.

Given the continued emergence of Omicron sub-lineages with additional immune escape mutations, we next modelled the impact of immune evasion on the stability of protection against symptomatic reinfection in a population that gained hybrid immunity by Omicron BA.1 or BA.2 breakthrough infection. Notably, even at timepoints where plasma neutralisation activity is maximal (∼1 month post-symptom onset), a novel variant with 3- or 10-fold reduction in neutralisation would reduce the protective efficacy from 90.1% to 74.1% or 46.8% respectively for BA.1 hybrid immune individuals, and from 95.3% to 85.5% or 63.4% respectively for BA.2 (Figure 4C, 4D). These fold change estimates are highly relevant, as comparable reductions in neutralisation activity have already been observed with recent Omicron sublineages BQ.1.1 and XBB (*10*). Overall, our modelling suggests that recovery from an Omicron breakthrough infection confers remarkably durable protection against closely related viral strains, but this protection can be rapidly subverted by the emergence of novel variants with increasing escape from neutralising antibodies.

## Discussion

The marked immune evasiveness and transmissibility of Omicron strains are driving increasing numbers of vaccine breakthrough infections worldwide, although the durability of immunity elicited by such infections remain unclear. Using intensive longitudinal sampling, we find both BA.1 or BA.2 breakthrough infection drive expansion of neutralising responses against the infecting strains from low or undetectable levels to high titres that were stably maintained from around 1 month post-symptom onset. Importantly, we show that only cross-reactive memory B cells were expanded by breakthrough infection, and the resulting antibody response was dominated by antibodies cross-reactive with ancestral spike, indicating that limited *de novo* responses were generated against neo-epitopes within BA.1 or BA.2 spike.

In line with recent studies (*3, 11*), our results are suggestive of immune imprinting, with no evident increase in BA.1 or BA.2 monospecific B cells even up to 4-7 months post-infection. Interestingly, Kaku et al. reported that BA.1 breakthrough infection drove affinity maturation of cross-reactive B cells and antibodies towards Omicron BA.1 over time (*11*), although our data suggests mechanistically this occurs through selective re-expansion of B cell memory. While the isolation of receptor binding domain (RBD)-specific monoclonal antibodies (mAbs) specific for the BA.1 RBD that do not cross-react with ancestral RBD has been reported, these comprised only a small fraction (median 4%) of the response to RBD (*11*), confirming that neo-epitopes are poorly recognised during breakthrough infection. Immune imprinting is not constrained to breakthrough infections, as monovalent Omicron BA.1 or bivalent Beta/Delta mRNA vaccines also predominantly boost pre-existing cross-reactive responses (*12*).

While superficially attractive, “overcoming” immune imprinting to generate responses against neo-epitopes may not actually be favourable for protection. Primary infection with Omicron elicits very limited neutralising breadth against historical non-Omicron variants (*13*). Similarly, Omicron-specific mAbs isolated from BA.1, BA.2 or BA.5 breakthrough individuals that do not bind ancestral RBD exhibit very narrow breadth of recognition, with little to no neutralisation activity against more recent Omicron variants including BQ.1.1 and XBB (*10*).

It remains unclear if there is something unique about “hybrid immunity” (vaccination then infection) in terms of durable reprogramming of the antibody response. Epidemiological studies have indicated that hybrid immunity confers stronger and more durable protection against infection compared to vaccine- or infection-elicited immunity alone (*14-17*). In line with other reports (*6, 18*), our data suggest that BA.1 and BA.2 breakthrough infection confers neutralisation breadth that extend to the more immune evasive BA.4 variant. Studies of a third dose of an ancestral spike vaccine have similarly demonstrated expansion of Omicron-specific B cells and enhancement of neutralisation breadth (*19, 20*), although not to the same extent as breakthrough infection with BA.1 (*6*). It remains to be seen whether the bivalent ancestral + BA.1 or ancestral + BA.5 mRNA vaccines can elicit equivalent levels of neutralisation breadth to Omicron breakthrough infection. Intriguingly, Muik et al. showed that neutralisation of BA.2 and BA.4/5 mediated by BA.2 breakthrough sera is highly dependent on antibodies against the N-terminal domain (NTD) of spike, while BA.1 breakthrough sera-mediated neutralisation is almost exclusively dependent on RBD antibodies (*21*). The different antibody targets elicited by BA.1 vs BA.2 breakthrough infection may have implications for immunity towards future Omicron variants, depending on which region of spike (RBD and/or NTD) gains further escape mutations (*10, 22*).

Both waning immunity and immune escape of new variants contribute to susceptibility of SARS-CoV-2 reinfection, though the relative contribution of these two factors is unknown. Here, we modelled the level of immune boosting and rate of antibody waning after breakthrough infection with BA.1 and BA.2 variants. Our analysis suggests robust and prolonged immunity to the homologous strain (70% protective efficacy for 4.5 and 7 years following BA.1 or BA.2 breakthrough infection). However, this protection is rapidly undermined by the emergence of more escaped variants. For example, a variant with a 3-fold reduction in neutralising antibody titre would reduce protective efficacy by the same amount expected after 3.8 years of waning immunity. A novel variant with a 10-fold reduction in neutralising antibody titre would reduce protective efficacy to 46.8% and 63.4% immediately following BA.1 or BA.2 breakthrough infection. This suggests that waning immunity likely plays a relatively small role in the ongoing susceptibility of SARS-CoV-2, especially with the emergence of BQ.1.1 (BA.5-derived) and XBB (BA.2-derived) variants that have displayed 9-37 fold drops in neutralisation relative to the homologous infecting strains in BA.1/BA.2 breakthrough cohorts (*10, 23, 24*).

Repeated, sequential waves of Omicron outbreaks have driven significant breakthrough infections and conferred “hybrid” immunity to much of the global population. Our data suggests that continuing spread of SARS-CoV-2 in this immune landscape will be less a function of waning immunity and in large part depend on the viral acquisition of further neutralisation escape mutations.

## Materials and methods

### Human subjects and ethics

A cohort of previously vaccinated participants with breakthrough COVID-19 (either PCR or rapid antigen test positive) were recruited through contacts with the investigators and invited to provide serial blood and nasal swab samples following symptom onset (Table S1), some of whom were previously described (*5*). Infecting variant was confirmed via whole genome sequencing of nasal swabs. For three participants where early nasal swabs were not available, the infecting variant was assigned based on predominant circulating strain at the time of infection. For all participants, whole blood was collected with sodium heparin anticoagulant. Plasma was collected and stored at -80ºC, and PBMCs were isolated via Ficoll-Paque separation, cryopreserved in 10% DMSO/FCS and stored in liquid nitrogen. Study protocols were approved by the University of Melbourne Human Research Ethics Committee (2021-21198-15398-3, 2056689), and all associated procedures were carried out in accordance with approved guidelines. All participants provided written informed consent in accordance with the Declaration of Helsinki.

### Analysis of viral RNA load by qPCR

For viral RNA extraction, 200 μL of nasal swab sample was extracted with the QIAamp 96 Virus QIAcube HT kit (Qiagen, Germany) on the QIAcube HT System (Qiagen) according to manufacturer’s instructions. Purified nucleic acid was then immediately converted to cDNA by reverse transcription with random hexamers using the SensiFAST cDNA Synthesis Kit (Bioline Reagents, UK) according to manufacturer’s instructions. cDNA was used immediately in the rRT-PCR or stored at -20ºC. Three microlitres of cDNA was added to a commercial real-time PCR master mix (PrecisionFast qPCR Master Mix; Primer Design, UK) in a 20 μL reaction mix containing primers and probe with a final concentration of 0.8μM and 0.1μM for each primer and the probe, respectively. Samples were tested for the presence of SARS-CoV-2 nucleocapsid (N) genes using previously described primers and probes (*25, 26*). Thermal cycling and rRT-PCR analyses for all assays were performed on the ABI 7500 FAST real-time PCR system (Applied Biosystems, USA) with the following thermal cycling profile: 95ºC for 2 min, followed by 45 PCR cycles of 95ºC for 5 s and 60ºC for 30 s for N gene.

### ELISA (N IgG and blocking ELISA)

Antibody binding to SARS-CoV-2 N protein was tested by ELISA. 96-well Maxisorp plates (Thermo Fisher) were coated overnight at 4°C with 2μg/mL recombinant N. After blocking with 1% FCS in PBS, duplicate wells of serially diluted plasma were added and incubated for two hours at room temperature. Bound antibody was detected using 1:20,000 dilution of HRP-conjugated anti-human IgG (Sigma) and plates developed using TMB substrate (Sigma), stopped using sulphuric acid and read at 450nm. Endpoint titres were calculated using Graphpad Prism as the reciprocal serum dilution giving signal 2× background using a fitted curve (4 parameter log regression).

For blocking ELISA, plates were coated overnight with recombinant BA.1 or BA.2 spike proteins (Hexapro ectodomain (*27*)). Plasma samples were pre-incubated in 20μl PBS with 1μg of ancestral spike protein, or BSA control for 1 hour prior to serial dilution in PBS+1%FCS containing 5μg/ml ancestral spike protein or BSA. Diluted plasma was added to the coated plate and incubated at room temperature for 30min, before washing and developing as described above.

### SARS-CoV-2 virus propagation and titration

Ancestral SARS-CoV-2 (VIC01) isolate was grown in Vero cells in serum-free DMEM with 1μg/ml TPCK trypsin while Omicron BA.1, BA.2 and BA.4 strains were grown in Calu3 cells in DMEM with 2% FCS. Cell culture supernatants containing infectious virus were harvested on Day 3 for VIC01 and Day 4 for Omicron strains, clarified via centrifugation, filtered through a 0.45μM cellulose acetate filter and stored at -80ºC. Infectivity of virus stocks was then determined by titration on HAT-24 cells (a clone of transduced HEK293T cells stably expressing human ACE2 and TMPRSS2 (*28*)). In a 96-well flat bottom plate, virus stocks were serially diluted five-fold (1:5-1:78,125) in DMEM with 5% FCS, added with 30,000 freshly trypsinised HAT-24 cells per well and incubated at 37ºC. After 46 hours, 10μl of alamarBlue™ Cell Viability Reagent (ThermoFisher) was added into each well and incubated at 37ºC for 1 hour. The reaction was then stopped with 1% SDS and read on a FLUOstar Omega plate reader (excitation wavelength 560nm, emission wavelength 590nm). The relative fluorescent units (RFU) measured were used to calculate %viability (‘sample’ ÷ ‘no virus control’ × 100), which was then plotted as a sigmoidal dose response curve on Graphpad Prism to obtain the virus dilution that induces 50% cell death (50% infectious dose; ID_50_). Each virus was titrated in quintuplicate in three independent experiments to obtain mean ID_50_ values.

### SARS-CoV-2 microneutralisation assay with viability dye readout

In 96-well flat bottom plates, heat-inactivated plasma samples were diluted 2.5-fold (1:20-1:12,207) in duplicate and incubated with SARS-CoV-2 virus at a final concentration of 2× ID_50_ at 37ºC for 1 hour. Next, 30,000 freshly trypsinised HAT-24 cells in DMEM with 5% FCS were added and incubated at 37ºC. ‘Cells only’ and ‘Virus+Cells’ controls were included to represent 0% and 100% infectivity respectively. After 46 hours, 10μl of alamarBlue™ Cell Viability Reagent (ThermoFisher) was added into each well and incubated at 37ºC for 1 hour. The reaction was then stopped with 1% SDS and read on a FLUOstar Omega plate reader (excitation wavelength 560nm, emission wavelength 590nm). The relative fluorescent units (RFU) measured were used to calculate %neutralisation with the following formula: (‘Sample’ – ‘Virus+Cells’) ÷ (‘Cells only’ – ‘Virus+Cells’) × 100. IC_50_ values were determined using four-parameter non-linear regression in GraphPad Prism with curve fits constrained to have a minimum of 0% and maximum of 100% neutralisation.

### Flow cytometric detection of SARS-CoV-2 spike-reactive B cells

Biotinylated recombinant SARS-CoV-2 Spike of ancestral Wuhan Hu-1 and Omicron (BA.1 or BA.2) strains were conjugated to streptavidin-APC or -PE fluorophores, respectively. PBMCs were thawed and stained with Aqua viability dye (Thermo Fisher Scientific) and then surface stained with Spike probes, CD19 ECD (J3-119) (Beckman Coulter), IgA VioBlue (IS11-8E10) IgM BUV395 (G20-127), IgD PE-Cy7 (IA6-2), IgG BV786 (G18-145), CD21 BUV737 (B-ly4), CD27 BV605 (O323), streptavidin BV510 (BD Biosciences), CD14 BV510 (M5E2), CD3 BV510 (OKT3), CD8a BV510 (RPA-T8), CD16 BV510 (3G8), and CD10 BV510 (HI10a) (BioLegend). Cells were washed twice with PBS containing 1% FCS and fixed with 1% formaldehyde (Polysciences) and acquired on a BD LSR Fortessa using BD FACS Diva.

### Modelling the kinetics of antibody recall and decay

We used a piecewise model to estimate the activation time and growth rate of various immune responses (neutralization and N IgG responses) following breakthrough infections. The model of the immune response *y* for subject at *i* time *y*_*i*_ can be written as:

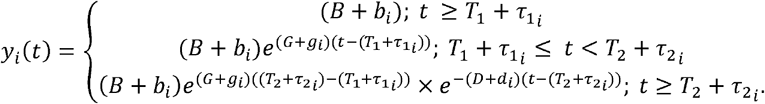

The model has 5 parameters; *B, G, T*_1_, *D* and *T*_2_. For a period before *T*_1_, we assumed a constant baseline value *B* for the immune response will grow at a rate of *G* until *T* _2_. From *T*_2_, the immune response a constant baseline value for the immune response. After the activation time *T*_1_, will decay at a rate of *D*. For each subject *i*, the parameters were taken from a normal distribution, with each parameter having its own mean (fixed effect). A diagonal random effect structure was used, where we assumed there was no correlation within the random effects. The model was fitted to the log-transformed data values, with a constant error model distributed around zero with a standard deviation σ. To account for the values less than the limit of detection, a censored mixed effect regression was used to fit the model. A categorical covariate was used to quantify the difference in parameters between different groups (i.e. BA.1 vs BA.2 group), and significance was determined based on the value of this binary covariate using a Wald test. Model fitting was performed using MonolixR2019b.

The decay rate of neutralisation was estimated by fitting a linear mixed effect model for each response variable as a function of days post-symptom onset, infection wave (i.e. BA.1 and BA.2), and response type (WT, matched, and BA.4). Likelihood ratio test was used to determine if the decay rate is different with respect to infection wave and response type. We fitted the model to log-transformed data of various response variables (assuming exponential decay), and we censored the data from below if it was less than the threshold for detection. The model was fitted by using *lmec* library in *R (v4*.*2*.*1)*, using the maximum likelihood algorithm to fit for the fixed effects.

### Predicting vaccine efficacy to different variants after breakthrough infection

We used the previously published model for predicting vaccine efficacy for a given plasma neutralisation titre (*9*). This model relates the neutralisation titre normalised to the geometric mean neutralisation titre against ancestral virus, of a convalescent cohort (individuals infected with the ancestral virus), with vaccine efficacy. Thus, we first estimated the geometric mean peak neutralisation titre for BA.1 and BA.2 breakthrough infections against three variants (the ancestral virus, the matched breakthrough variant and BA.4), and normalised these to a previously reported cohort (n=8) of convalescent individuals (*29*) using the same neutralisation assay (Geometric mean neutralisation titre of this cohort 507.6). Pooling the data on the decay of neutralisation across all individuals and variants we estimated the kinetics of neutralisation against each of these variants up to 3650 days after the peak. These predicted kinetics of neutralisation titres were then used in the model by Khoury et al. to predict vaccine efficacy at each neutralisation titre using the formula:

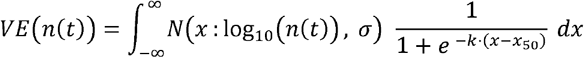

Where *N (x* : *μ, α)* is the probability density function of a normal distribution with mean μ and standard deviation α, σ is the standard deviation of log_10_ neutralisation titres of the vaccinated population, *n(t)* is the geometric mean neutralisation titre at time *t* days after the peak neutralisation titre (normalised to the geometric mean of the convalescent cohort), and *X*_50_ is the log_10_ neutralisation titre associated with 50% protection. The parameters σ, *K* and *X*_50_ were estimated in Khoury et al. along with standard errors of the parameter estimates (*9*). Confidence intervals of the predictions were estimated by parametric bootstrapping as described previously (*30*). In brief, 10,000 random samples of the estimated peak neutralisation titre, decay rate, and model parameters σ, *k* and *X*_50_ were randomly drawn from normal distributions around the mean estimates of each parameter using the standard error of each estimate or covariance matrix where appropriate, the VE was estimated using the above equation for each set of sampled parameters and the 2.5^th^ and 97.5^th^ percentiles of the bootstrapped estimates at each neutralisation titre were calculated as the lower and upper 95% confidence bounds, respectively (indicated by shaded region in figure 4).

## Data Availability

All data produced in the present study are available upon reasonable request to the authors

## Acknowledgements

We thank the participants for the generous involvement and provision of samples. We thank Grace Gare and Andrew Kelly (University of Melbourne) for excellent technical assistance. We thank molecular staff at the Victorian Infectious Diseases Reference Laboratory for performing RT-PCR. We thank Dr Julian Druce and Dr Leon Caly at the Victorian Infectious Diseases Reference Laboratory for isolating and distributing SARS-CoV-2 virus isolates. We acknowledge the Melbourne Cytometry Platform for provision of flow cytometry services.

## Author contributions

Conceptualization: WSL, HXT, JAJ, SJK, AKW

Formal Analysis: WSL, HXT, AR, DC, MPD, DSK, AKW

Funding Acquisition: MPD, SJK, AKW

Investigation: WSL, HXT, RE, MK, JN, TA, HEK, GT, PK, KCL, TT, AKW

Methodology: WSL, HXT, AR, AA, SGT, DSK, AKW

Resources: GT, PK, KCL, TT, AA, SGT, DAW

Supervision: DC, MPD, SJK, JAJ, DSK, AKW

Writing – original draft: WSL, HXT, AKW

Writing – review & editing: WSL, HXT, RE, MK, GT, DAW, MPD, SJK, JAJ, DSK, AKW

## Funding

Australian National Health and Medical Research Council grants 1149990, 1162760 and 2004398

Australian Medical Research Future Fund grants 2005544 and 2013870

The Victorian Government

Australian National Health and Medical Research Council Investigator or Fellowship grants (HXT, MK, DAW, MPD, SJK, JAJ, AKW)

Melbourne Postdoctoral Fellowship (WSL)

## Competing interests

The authors declare no competing interests.

## Data and materials availability

All data are available in the main text or the supplementary materials.

## Supplementary material

**Figure S1.**
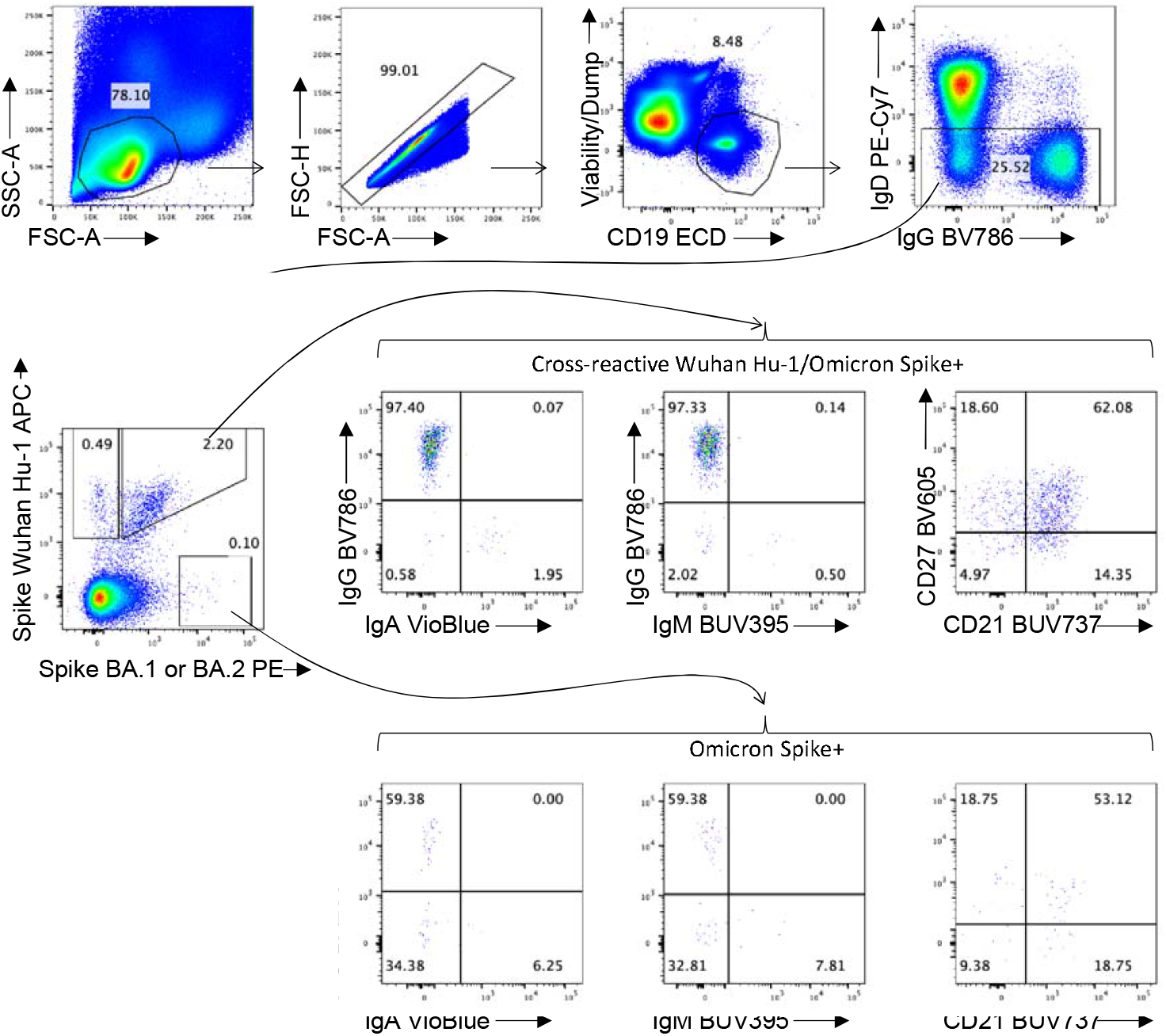
Gating strategy for the detection and phenotyping of spike-specific B cells. Lymphocytes were identified by FSC-A vs SSC-A gating, followed by doublet exclusion (FSC-A vs FSC-H), and gating on live CD19+ B cells. Class-switched B cells were identified as IgD-. Binding to SARS-CoV-2 ancestral Wuhan Hu-1 or Omicron (BA.1 or BA.2) spike was assessed. Cross-reactive (Wuhan Hu-1+ Omicron+) or mono-specific (Omicron+) B cells were assessed for surface IgM, IgG or IgA isotypes, and CD21 and CD27 co-expression.

**Supplementary Table 1.** Breakthrough infection cohort demographics

| Subject | Gender | Breakthrough infection strain | Vaccination/infection history | No. of prior vaccinations and/or infections | Last vaccine to symptom onset (days) | Acute longitudinal sampling (Fig 1B,1C, 2A) | MBC analysis (Fig 3) | Decay analysis (Fig 2C) |
| --- | --- | --- | --- | --- | --- | --- | --- | --- |
| COR015 | F | BA.1 | 3x BNT162b2 | 3 | 38 | □□ | □□ | □□ |
| COR032 | F | BA.1 | 2x ChAdOx1 nCoV-19, 1x mRNA-1273 | 3 | 32 | □□ | □□ | □□ |
| CP110 | M | BA.1 | 2x BNT162b2 | 2 | 84 | □□ | □□ | □□ |
| CP111 | M | BA.1 | 2x BNT162b2 | 2 | 100 | □□ | □□ |  |
| CP112 | F | BA.1 | 2x BNT162b2 | 2 | 90 | □□ |  |  |
| COR198 | F | BA.1 | 2x ChAdOx1 nCoV-19, 1x mRNA-1273 | 3 | 39 | □□ | □□ | □□ |
| CP069 | F | BA.1 | Infection (Wuhan), 2x BNT162b2 | 3 | 111 | □□ | □□ | □□ |
| COR005 | M | BA.1 | 1x NVX-CoV2373, 2x BNT162b2 | 3 | 102 | □□ | □□ | □□ |
| COR012 | F | BA.1* | 3x BNT162b2 | 3 | 38 |  |  | □□ |
| COR036 | F | BA.2 | 2x BNT162b2, 1x mRNA-1273 | 3 | 90 | □□ | □□ |  |
| COR274 | F | BA.2 | 2x ChAdOx1 nCoV-19, 1x mRNA-1273 | 3 | 155 | □□ | □□ | □□ |
| COR275 | M | BA.2 | 2x ChAdOx1 nCoV-19, 1x mRNA-1273 | 3 | 83 | □□ | □□ | □□ |
| COR291 | M | BA.2 | 3x BNT162b2 | 3 | 124 | □□ | □□ |  |
| COR039 | F | BA.2 | 2x BNT162b2, 1 x mRNA-1273 | 3 | 169 | □□ |  | □□ |
| COR215 | F | BA.2 | 2x ChAdOx1 nCoV-19, 1x mRNA-1273 | 3 | 125 | □□ | □□ | □□ |
| COR216 | M | BA.2 | 2x ChAdOx1 nCoV-19, | 3 | 122 | □□ |  |  |
|  |  |  | 1x mRNA-1273 |  |  |  |  |  |
| CP120 | F | BA.2 | 3x BNT162b2 | 3 | 151 | □□ |  |  |
| COR043 | F | BA.2 | 2x ChAdOx1 nCoV-19,<br>1x mRNA-1273 | 3 | 64 | □□ | □□ | □□ |
| CP116 | M | BA.2 | 2x ChAdOx1 nCoV-19,<br>1x mRNA-1273 | 3 | 85 | □□ | □□ |  |
| CP117 | M | BA.2 | 3x BNT162b2 | 3 | 52 | □□ | □□ | □□ |
| COR281 | F | BA.2 | 3x BNT162b2 | 3 | 118 | □□ | □□ |  |
| CP118 | M | BA.2 | 2x ChAdOx1 nCoV-19,<br>1x mRNA-1273 | 3 | 100 | □□ | □□ |  |
| CP119 | F | BA.2 | 2x BNT162b2,<br>1x mRNA-1273 | 3 | 86 | □□ | □□ |  |
| CP40 | M | BA.2 | Infection (Wuhan),<br>2x ChAdOx1 nCoV-19,<br>1x BNT162b2 | 4 | 49 | □□ | □□ |  |
| COR011 | F | BA.2* | 3x BNT162b2 | 3 | 125 |  |  | □□ |
| COR282 | M | BA.2* | 3x BNT162b2 | 3 | 157 |  |  | □□ |

\*Nasal swabs to determine infecting variant were unavailable, thus infecting strain was assigned by predominant circulating strain at the time of infection.

**Supplementary Table 2.** Piecewise linear regression parameters of N-binding IgG (with 95% CI) following BA.1 or BA.2 breakthrough infection. Related to Fig 1B.

| N IgG | Estimated parameter (95% CI) |  |  |
| --- | --- | --- | --- |
|  | BA.1 vs BA.2 |  |  |
|  | BA.1 | BA.2 | p-value<br>(for the difference) |
| Activation time (days post onset) | 7.4<br>(5.4 - 8.7) | 6.7<br>(3.4 - 9.6) | 0.63 |
| Doubling time (days) | 2.4<br>(1.9 - 3.7) | 1.5<br>(0.9 - 4) | 0.08 |
| Fold change (from baseline) | 9<br>(1.6 - 16.5) | 14.5<br>(7.9 - 21) | 0.24 |

**Supplementary Table 3.** Piecewise linear regression parameters of neutralising antibodies (with 95% CI) following BA.1 or BA.2 breakthrough infection. Related to Fig 2A.

| Neutralization<br>IC<sub>50</sub> | Estimated parameter (95% CI) |  |  |  |  |  |  |  |  |
| --- | --- | --- | --- | --- | --- | --- | --- | --- | --- |
|  | BA.1 breakthrough infection |  |  | BA.2 breakthrough infection |  |  | BA.1 vs BA.2 |  |  |
|  | WT | BA.1 | p-value<br>(for the<br>difference) | WT | BA.2 | p-value<br>(for the<br>difference) | BA.1 | BA.2 | p-value<br>(for the<br>difference) |
| Activation<br>time (days<br>post onset) | 2.3<br>(0.3 - 20.3) | 3.1<br>(1.5 - 6.6) | 0.82 | 2.3<br>(0.8 - 7.2) | 3.6<br>(2.4 - 5.6) | 0.55 | 3.1<br>(1.5 - 6.6) | 3.6<br>(2.4 - 5.6) | 0.78 |
| Doubling time<br>(days) | 6.1<br>(3.3 - 11.3) | 2.1<br>(1.5 - 3) | 0.02 | 7.8<br>(5.4 - 11.2) | 2.8<br>(2.3 - 3.5) | 0.000013 | 2.1<br>(1.5 - 3) | 2.8<br>(2.3 - 3.5) | 0.26 |
| Fold change<br>(from<br>baseline) | 5.4<br>(2.2 - 8.7) | 31<br>(13.7 - 48.3) | 0.0078 | 15.6<br>(6.5 - 37.8) | 34.7<br>(11.9 - 57.4) | 0.0042 | 31<br>(13.7 - 48.3) | 34.7<br>(11.9 - 57.4) | 0.53 |

**Supplementary Table 4.** Decay rates of neutralising antibodies (with 95% CI) following BA.1 or BA.2 breakthrough infection. Related to Fig 2C.

| Neutralization<br>IC<sub>50</sub> | Estimated parameter (95% CI) |  |  |  |  |  |  |  |  |
| --- | --- | --- | --- | --- | --- | --- | --- | --- | --- |
|  | BA.1 breakthrough infection |  |  | BA.2 breakthrough infection |  |  | BA.1 vs BA.2 |  |  |
|  | WT | BA.1 | p-value<br>(for the<br>difference) | WT | BA.2 | p-value<br>(for the<br>difference) | BA.1 | BA.2 | p-value<br>(for the<br>difference) |
| Half-life<br>(days)* | 182.8<br>(110.7 - 523.8) | 333.5<br>(136.9 - 10000) | 0.98 | 1216<br>(252.6 - 10000) | 1049.7<br>(210.5 - 10000) | 0.43 | 333.5<br>(136.9 - 10000) | 1049.7<br>(210.5 - 10000) | 0.36 |

\*half-life upper limit is set to 10000 days

## Notes

### Competing Interest Statement

The authors have declared no competing interest.

### Funding Statement

 This study was funded by:
- Australian National Health and Medical Research Council grants 1149990, 1162760 and 2004398
- Australian Medical Research Future Fund grants 2005544 and 2013870
- The Victorian Government
- Australian National Health and Medical Research Council Investigator or Fellowship grants (HXT, MK, DAW, MPD, SJK, JAJ, AKW)
- Melbourne Postdoctoral Fellowship (WSL)

### Author Declarations

Ethics committee/IRB of University of Melbourne Human Research Ethics Committee gave ethical approval for this work. Study protocols were approved by the University of Melbourne Human Research Ethics Committee (2021-21198-15398-3, 2056689), and all associated procedures were carried out in accordance with approved guidelines. All participants provided written informed consent in accordance with the Declaration of Helsinki.

